# A Threshold Criteria for Seasonal Amplification and Outbreaks of Mosquito-Borne Disease (MBD) Cases in Kerala using Climate Parameters

**DOI:** 10.1101/2022.11.10.22282112

**Authors:** Rajib Chattopadhyay, Divya Surendran, S Lekshmi, Pulak Guhathakurta, K.S. Hosaliker, D.S. Pai, M. S Manu, M. Mohapatra

## Abstract

Modelling the dynamics of mosquito borne disease (MBD) cases is a challenging task. The current study first proposes a generic dynamical model to qualitatively understand the seasonality as well as outbreaks of malaria and dengue over the state of Kerala based on a climate forced oscillator model, which is then supplemented by a data driven model for quantitative evaluation. The proposed forced oscillator model is parametric and general in nature which can be qualitatively used to understand the seasonality and outbreaks. However, since parametric model-based estimation require estimation of multiple parameters and several closure assumptions, we used the K-means clustering which is a data driven clustering approach to understand the relationship between Malaria and Dengue cases and climate forcing. The results showed a clear relationship of the MBD cases with the first order and second order moments (i.e. mean and standard deviation) of the climate forcing parameters. Based on this, we came up with an objective threshold criterion which relates the climate parameters to the number of cases of malaria and dengue cases over Kerala.

## Introduction

Temporal evolution of Mosquito borne disease (MBD) like malaria and dengue are known to be related to local tropical humid climatic conditions. India report a large number of MBD every year. These MBD cases show rapid seasonal peaks and sometimes cause severe localized outbreaks leading to several fatalities. In the recent years, after the adoption of National Framework for Malaria Elimination (NFME) and the National Strategic Plan for Malaria Elimination (2017–22), there is a significant decrease in malaria cases in India(Narain and Nath 2018; Mohan et al. 2021; NVDCP,,MoHFW 2022). However, local outbreaks of MBD like malaria and dengue is commonly reported over different states of India and provide several unique challenges for detection and elimination (Das et al. 2012; Gupta et al. 2012; Singh et al. 2020; Ranjha and Sharma 2021; Paradkar et al. 2021). As per WHO world malaria report 2021, India contributed 1.7% of malaria cases and 1.2% deaths globally during 2021 (WHO 2021). Similarly, WHO reports that dengue spread over 128 countries across the globe and affected ∼5 million people during the year 2020.

India shows different state/regional gradation in terms of number of malaria and dengue cases(Das et al. 2012; Gupta et al. 2012; Singh Parihar et al. 2019). Prevention of such localized VBD outbreaks like malaria and dengue and timely intervention to reduce the disease burden is important from community health practice perspective. Epidemiological modelling and simulation studies often relate the vector growth and proliferation dynamics to the ambient weather and climatic factors like temperature, humidity, rainfall etc. (Loevinsohn 1994; Craig et al. 1999; Paaijmans et al. 2009; Laneri et al. 2010; Hii et al. 2012; Lunde et al. 2013; Singh Parihar et al. 2019; Patil and Pandya 2021; Colón-González et al. 2021). The studies showed that temporal evolution of these climate factors provides necessary “forcing” for temporal evolution dynamics. Several studies also assume the environmental forcing as a driving factor for a near future estimation/outlooks of such outbreaks using linear, non-linear, dynamical as well as statistical forecasting models (Laneri et al. 2010; Hii et al. 2012; Zinszer et al. 2015; Anwar et al. 2016; Sewe et al. 2017; Hussien 2019; Patil and Pandya 2021; Nkiruka et al. 2021). Although, epidemiological model of malaria and dengue outbreaks relates several external factors to the growth of the parasite/mosquito, temperature and other climate derived factors are known to contribute to the growth of both the malarial parasite and mosquito. Hence, such studies conclude that climatic factors are extremely important for MBD outbreaks. In the context of climate change scenario and rise in global mean temperature, studies reports frequent proliferation of the VBD outbreaks(Reiter P 2001; Caminade et al. 2014; Campbell et al. 2015; Ebi and Nealon 2016; Colón-González et al. 2021).

How do the climatic factors are related to seasonal evolution of MBD and its occasional outbreaks i.e. increase in large number of cases over a particular location for a short period of time? Do climate extremes force such outbreaks? The climatic fields such as temperature, humidity, rainfall can be assumed to be the forcing, causing an increase or decrease in the mosquito population or creating favourable condition for the development of the malarial parasites. An Example is shown in **Fig.1a and 1b**, which shows the seasonal cycle (i.e. average month wise distribution) of the number of reported malaria and dengue cases for all districts of Kerala and accumulated rainfall over Kerala (**Fig.1c)** for the recent 12 years 2011-2021. The clear summer season peak of both the time series is evident for all years. Such seasonal peaks of malaria are reported in other states also and the monthly variations of malaria cases resembles the rainfall variations. The outbreaks above the seasonal cycles, when there are excess values in different months are also known to be related to climate factors in different regions (Laneri et al. 2010). The power spectral plot of the malaria, dengue along with rainfall and few other climatic variables for Kerala are shown in **Fig.2**. The plot shows an annual cycle and other statistically significant peaks (>90% confidence) for different climatic variables (spectral peaks above the green line is significant). **Fig.2** shows the similar frequency response of malaria and climate variables (Rainfall, maximum temperature (Tmax), minimum temperature (Tmin), Relative Humidity (RH) at 3UTC and at 12UTC) for the time period considered in this study. **Fig.1** and **2**, thus represents the similar cyclical variability for both the variables. Physically MBD seasonal evolution cycle can be linked to temperature or rainfall cycle as the parasite growth and development are favoured by climatological condition. In addition to such cyclical behaviour, how does the outbreak of disease (i.e. increase in the number of Malaria/Dengue cases for few months relative to the expected or mean value) relate to the climatic factors?

**Figure 1.**
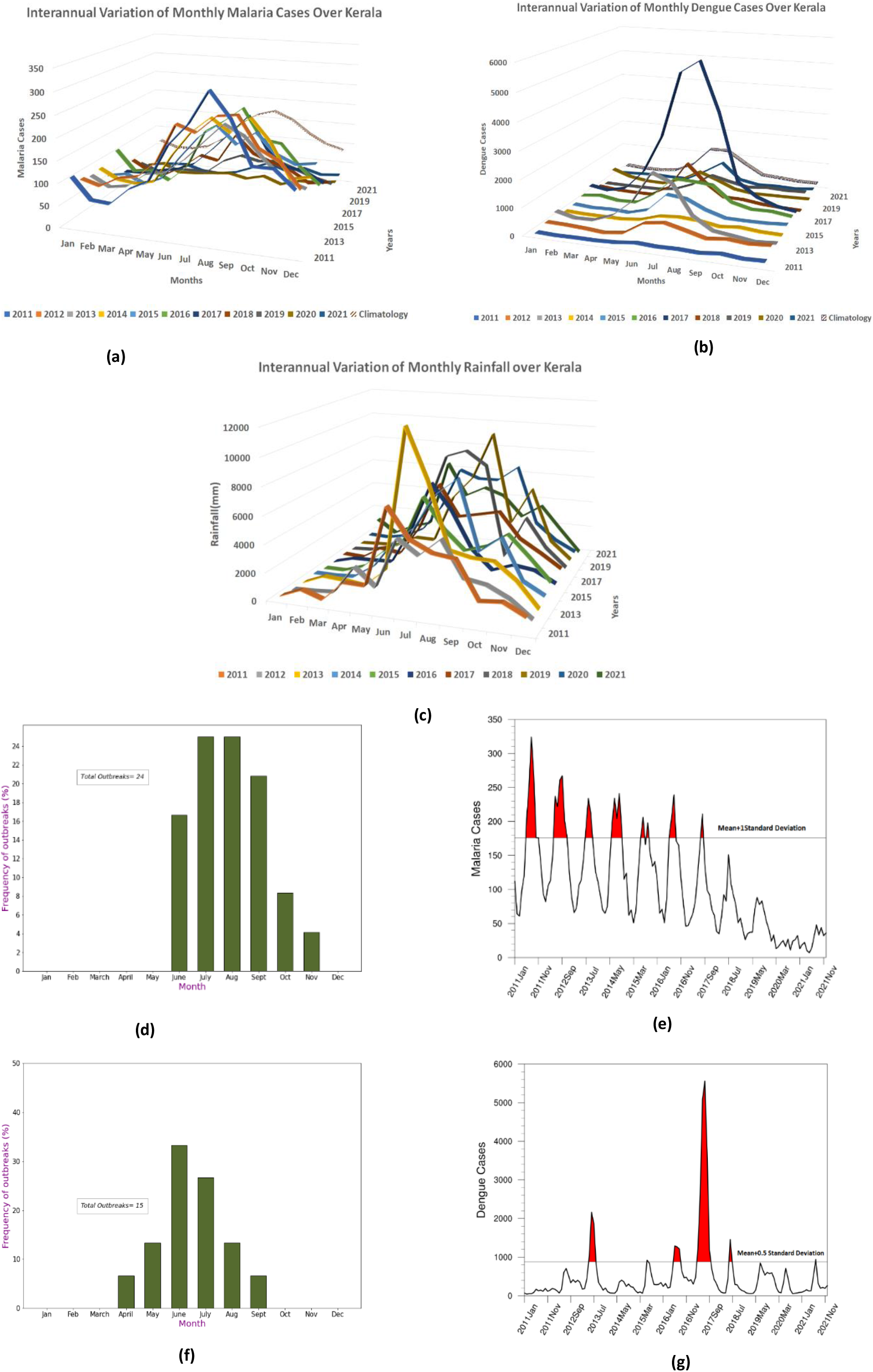
**(a)**. Interannual variation of monthly Malaria Cases over Kerala. **(b)**. Interannual variation of Dengue Cases over Kerala. **(c)**. Interannual variation of monthly Rainfall over Kerala. **(d)**. Monthly distribution of Outbreak frequency of Malaria Cases (Mean + 1 SD). **(e)**. Outbreak frequency of Malaria Cases (Mean + 1 SD). **(f)**. Outbreak frequency of Dengue Cases (Mean + 0.5 SD). **(g)**. Outbreak frequency of Dengue Cases (Mean + 0.5 SD).

**Figure 2a to 2f:**
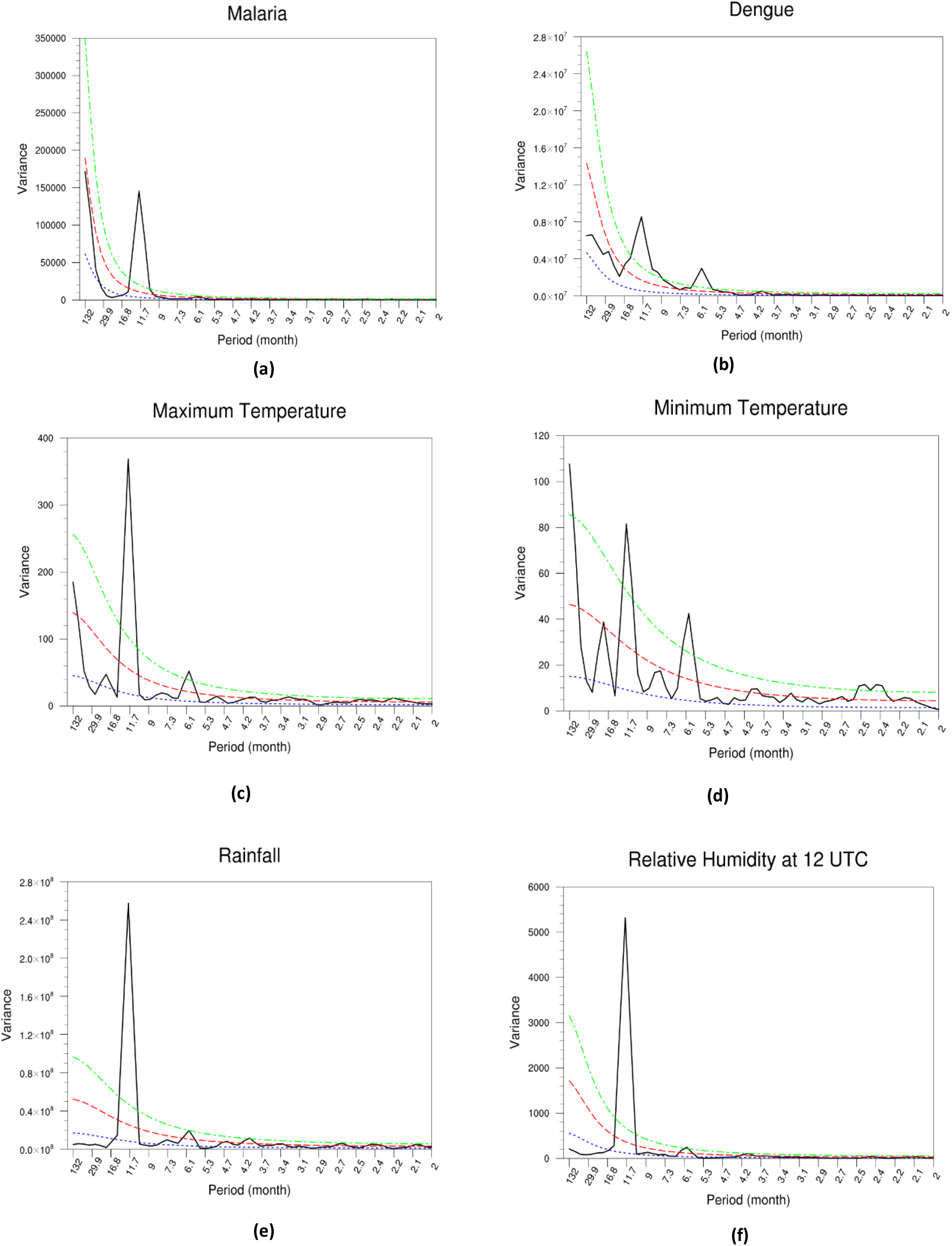
Power spectrum for Malaria, Dengue, Maximum Temperature, Minimum Temperature, Rainfall and Relative Humidity at 12 UTC.

Based on a generic model, the current study reports a case study based on the malaria and dengue data from Kerala and estimate the climatological conditions or thresholds related to the malaria and dengue outbreaks during the period 2011-2021. This generic model would first show the seasonal cycle evolution and then it will provide conditions when the outbreaks can be forced by climatological components. The generic model is shown in **Fig.3**. It is a coupled model relating the malaria/dengue cases with the vector number density. We assume that the vector density is coupled to climate factors rather than the reported number of malaria/dengue case. The dynamics of malaria or dengue cases (M(t)) can be given as:

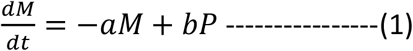

and, the number of vector (P(t)) forced by climate component (W) as:

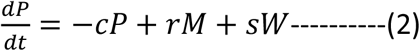

**Figure 3:**
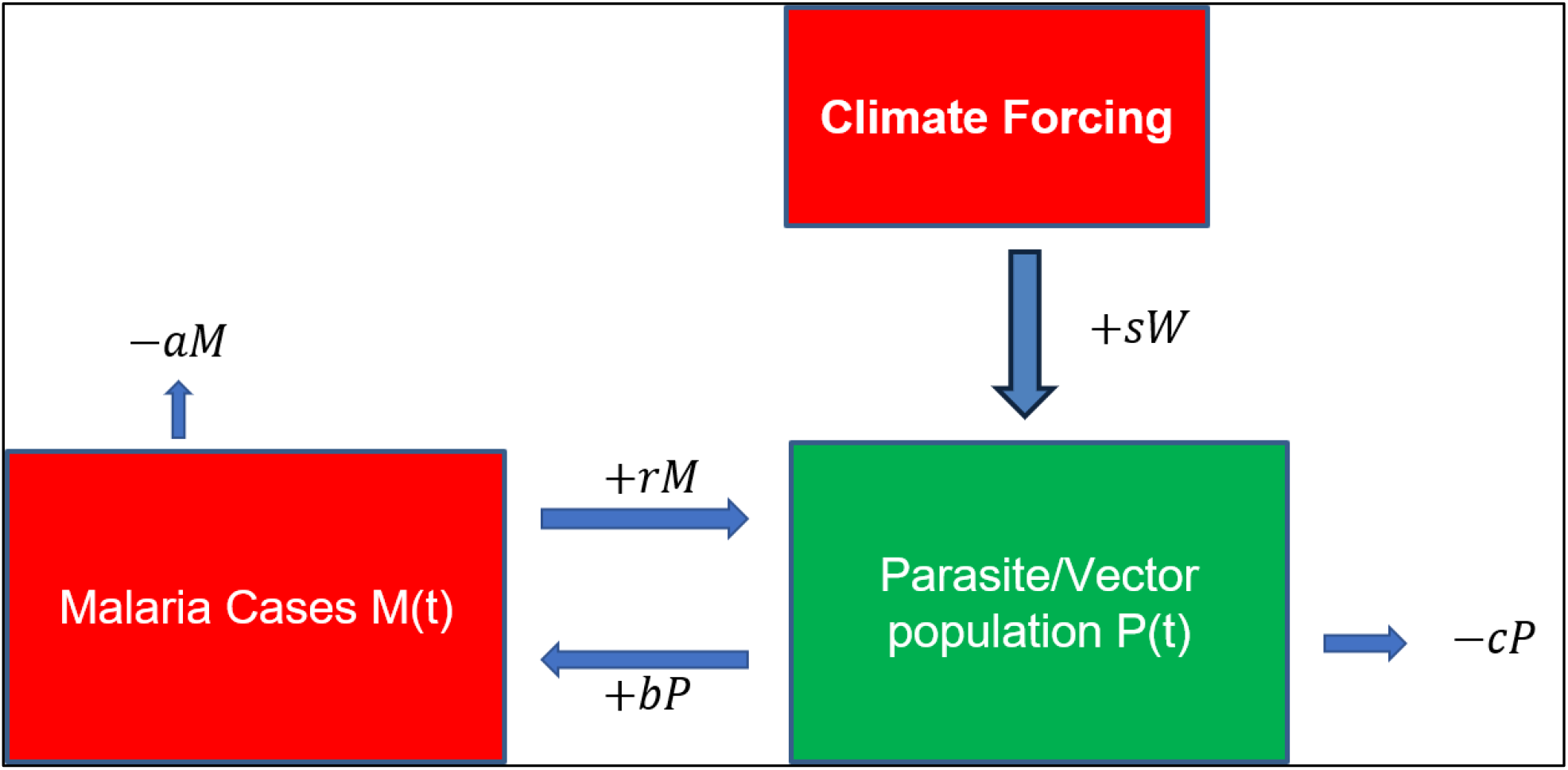
A schematic model of Malaria dynamics with Climate Forcing.

In the above equations, the sign of the constants (a,b,c…etc.) are given to represent sign of feedback. Negative feedbacks in an equation is represented by negative sign and the positive feedbacks are represented by positive sign for that equation. Thus, the rhs terms [−*aM*, +*bP*] in equation (1) represents the negative and positive feedbacks to 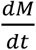 leading to decrease or increase of the lhs term. Similarly, for the temporal change of parasite number(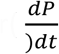 term, the positive and negative feedback terms are [−*cP, rM, sW*] constructed. [*sW*] represents the weather feedback term. Note that the weather feedback term impacts the parasite (i.e. equation 2), rather than the number of cases (equation 1). There could be other terms, but the simple coupled relationship is assumed for highlighting the impact of climatic factors. The medical interventions, genetic resistance against the MBD in the sample population are assumed in [-aM] or [-cP] terms. A combination of the terms leads to the following equation:

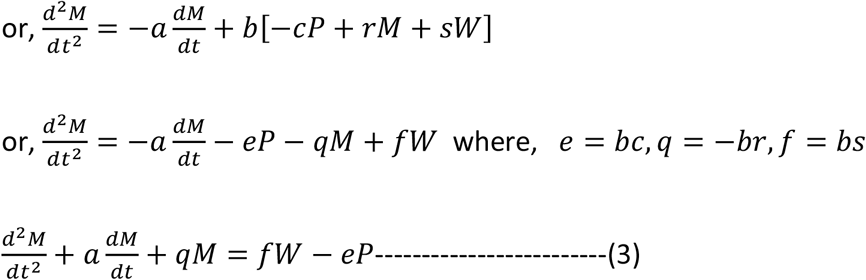

The above equation represents a damped oscillator with a forcing term *F = fW* − *eP* in the right-hand side. It is to be noted that the climate forcing is a function of multiple climate parameters, i.e. *W* ≡ *W(rainfall, temperature, humidity, etc.)*. Clearly the climate component and the MBD cases are a periodic function as shown in ***Fig.1*** and there can be two situation **(I)**, case when e=0, and more general when (II) *e* ≠ 0

**Case(I)**: If it is assumed that the parasitic forcing is negligible (due to control of mosquitos or other medical interventions), i.e. second term is close to zero, i.e. or e=0 (*e* ≠ 0 *in general)*, the equation 3 simplifies to:

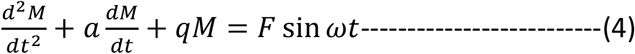

Assuming a periodic solution, *Re(Ae*^*i(ωt*+*φ)*^),the amplitude and phase is given by

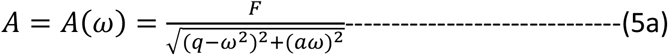

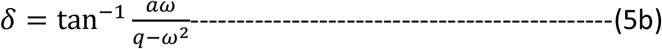

The solution of M(t) is given by

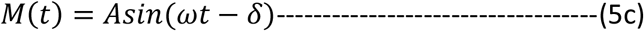

The above solutions suggest two things: (a) the climate forcing can drive the seasonal cycle; a periodic forced solution of M(t) following the climate forcing sinusoidal, relate MBD annual cyclical evolution to climate annual cycle as shown in **Fig.1**, and (b) the amplitude of the seasonal cycle, ***A*** depends on Climate forcing amplitude ***F*** and the denominator of (5a). **F** is in general a function of multiple climate variables (temperature, rainfall humidity etc), and the denominator of (5a) is minimum when *ω = q (resonant criteria)*. These two conditions as mentioned above defines the amplitude of M(t). Malaria/dengue outbreak or extreme values of ***M***(t), thus, is related to climate factors **F** and a resonant interaction. ‘

**Case (II)** when *e ≠* 0: This is the more general case. Since the first and the second term in the rhs side compete with each other (opposing sign), the net forcing is related to the temporal evolution of malarial parasite at any time. If the net forcing is zero, i.e. *fW* − *eP =* 0, the equation shows malaria cases decreasing with time (damped oscillator with no external forcing). This can be the case, for example when there is strong biological or medical intervention and climate action balance the parasite growth. The decreasing trend in malaria cases intuitively suggest such situation. However, this does not mean climate forcing has become less important. It simply means that the oscillator is not forced into action due to human intervention. In the context of climate change, if the climate forcing changes in the (**e,f**) parameter space, the cases would again increase. It may be also noticed that the second term on the right-hand side of Eq.3, it adds stochastic noise depending on the amplitude of the constants. Also, the amplitude of climate forcing can also be stochastic. So, it can be said that the outbreaks of the MBD above the cyclical variation can be related to this stochastic forcing. Thus equation (5) provides a basis for seasonal cycle and outbreak conditions both for malaria and any other MBD. It also emphasizes the importance of climate change. In a changed regime, climate forcing can in fact force the oscillator to increase the malaria cases.

The current study would not provide a mathematical solution for this outbreak conditions (i.e. estimate ***e*** and ***f***), rather it would explore under what climatological conditions (represented by a set of climate variables described earlier) the severe outbreaks of MBD (represented by number of reported cases) can occur in a non-parametric sense i.e. what climate forcing leads to larger malaria/dengue cases. Parametric models describing vector dynamics deals with large number of parameters(Taghikhani and Gumel 2018; Singh Parihar et al. 2019; Lee et al. 2021) and can be often sensitive to these parameters. The non-parametric relationship is simplistic and data driven and would be useful as it is found that there is linear as well as noise and non-linearity mixed relationship between malaria and several climate variables which imparts non-stationarity in the data and hence can be less reliable. In fact, the forcing is related to multiple climate variables as described earlier provides multiple source of uncertainties in parasite dynamical model (i.e. equation 2). This is shown in **Fig.4**. The variables are chosen based on previous studies (Guhathakurta et al. 2021; Patil and Pandya 2021; Dutta et al. 2021). It can be seen that many surface variables are correlated with the malaria and dengue cases. Hence, there is a relationship between these variables and MBD evolution and outbreaks. Thus, malaria/dengue outbreak could be traced by multivariate clustering analysis. Clustering analysis would identify the severity thresholds associated with the outbreaks. The severity threshold, thus identified, would help to derive large scale climate conditions for the disease outbreak and hence its prediction.

**Figure 4:**
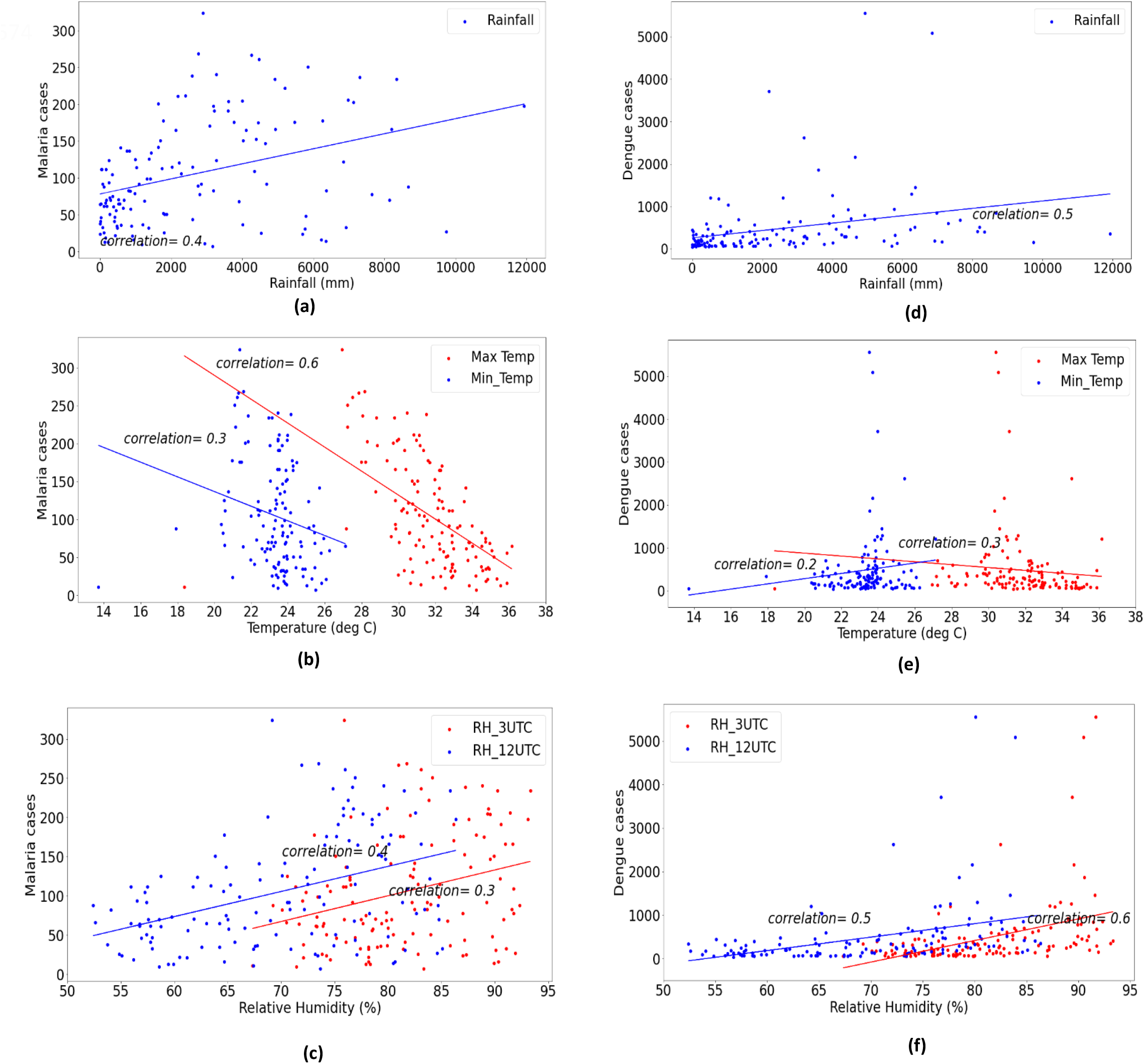
Correlation of Malaria and Dengue cases with different climate parameters. **(a)**. Malaria and Rainfall **(b)**. Malaria and Relative Humidity **(c)**. Malaria and Maximum & Minimum temperatures **(d)**. Dengue and Rainfall **(e)**. Dengue and Relative Humidity **(f)**. Dengue and Maximum & Minimum temperatures

## Data and Methods

### (i) Data

#### Disease Data

Monthly data of malaria and dengue cases over Kerala was collected from State News bulletin on Integrated Disease Surveillance Project (ISDP) for the years 2011 to 2021 (132 months). https://dhs.kerala.gov.in/data-on-communicable-diseases/

#### Meteorological Data

Climate variables (Rainfall, Tmax, Tmin and Relative Humidity at 3UTC & 12UTC) used for clustering analysis has been calculated from the station wise data of Kerala obtained from National Data Centre (NDC), Pune (https://dsp.imdpune.gov.in/).

For the analysis of the spatial evolution of the meteorological factors on the Malaria outbreak pattern, the composite of variables such as daily maximum temperature (Tmax), minimum temperature (Tmin), Skin (i.e earth’s surface) Temperature, relative humidity (RH) at 12 UTC and rainfall has been used. All the datasets except rainfall has been obtained from the NCEP-NCAR Renanalysis-1 dataset (Kalnay et al. 1996) with a spatial resolution of 2.50° x 2.50°. The monthly mean surface Tmax and Tmin are obtained directly from the reanalysis products. The daily mean skin temperature and the surface RH at 12 UTC are used to calculate the respective monthly mean values. Similarly, the daily gridded rainfall data of spatial resolution of 0.25° x 0.25° from India Meteorological Department (Pai et al. 2014) has been used to obtain the monthly mean rainfall (mm/day) and all the variables are analysed for the same period as above from 2011-2021 (132 months).

### (ii) Methods

Clustering is a form of unsupervised learning whereby a set of observations (i.e., data points) is partitioned into natural groupings or *clusters* of patterns in such a way that the measure of similarity between any pair of observations assigned to each cluster minimizes a specified cost function (Haykin 2009). K-Means clustering is one such method in which the Euclidean distance between the points is used to aggregate the points into different clusters with the target number of clusters (k) to be provided initially by the user. The imaginary or real location representing the centre of any cluster is termed as *centroid*. Initially, we have to define the number of clusters to be obtained (k). A random ‘k’ points or centroids are selected from the dataset and each data point is assigned to their closest centroid, which will form the predefined ‘k’ clusters. The k-Means algorithm computes the centroids and repeats until the optimal centroid is found. The data points are assigned to clusters in such a way that the sum of the squared distances between the datapoints and the centroid is as small as possible.

In this study, the monthly mean of six variables e.g. Tmax, Tmin, RH at 3 UTC and 12 UTC, (accumulated) precipitation in Kerala and the number of reported Malaria/Dengue cases in the state has been used to construct a vector space with the vector having six dimensions for every month. The 132 monthly values (for the years 2011-2021) of this vector is used to identify a set of 5 clusters using K-Means clustering algorithms using a standard scikit-learn python module (https://scikit-learn.org/stable/modules/generated/sklearn.cluster.KMeans.html). Here each month represent a point in the six-dimensional vector space and the points are used to calculate the Euclidean distance for obtaining the optimized centroid of each cluster. In general, the clustering analysis will help to aggregate these points into different clusters based on the hidden patterns in the observed datasets representing a relation between the climatic variables (Tmax, Tmin, skin temperature, relative humidity, precipitation) and the vector borne disease (malaria/dengue) data.

### (iii) Outbreak criteria

In this analysis we have used a simple criterion for deciding the outbreak (i.e. extreme event) criteria in a month. For malaria cases considered, we have defined outbreaks when the standardized anomaly of cases in a month is greater than 1 standard deviation. For dengue, we have used 0.5 standard deviation criteria. This criteria is not quantitive, is not used quantitatively in

## Results

### (i) Clustering results

Clustering for Malaria cases is shown in **Fig.5** from panel **(a)** to **(e)** for the clusters 1 to 5 along with multiple climate variables used in the clustering. The variables (represented as bars) of each cluster which are used for the clustering are plotted as a ratio of the corresponding time series mean. Suppose the Tmax in one cluster is 1.75, it means that the value of Tmax of that cluster is 1.75 times greater than the mean of Tmax time series. Similarly, the standard deviation is plotted as a fraction of the standard deviation of time series. In cluster2, which has a highlighted frame, the number of malaria cases are approximately two times greater than the malaria time series mean. It shows that cluster 2 has a greater number of malaria cases as compared to the other clusters. The climate variables in cluster 2 show some specific values; like rainfall is 1.75 times greater than the rainfall time series mean and the Tmax, Tmin and relative humidity are close to their corresponding time series mean. The mean and standard deviation of all variables in actual time series along with that for each malaria cluster are given in **Table-1**. So, it means that these specific combinations given in **Table-1** for the cluster 2 have the potential to bring more malaria cases.

**Table-1.**
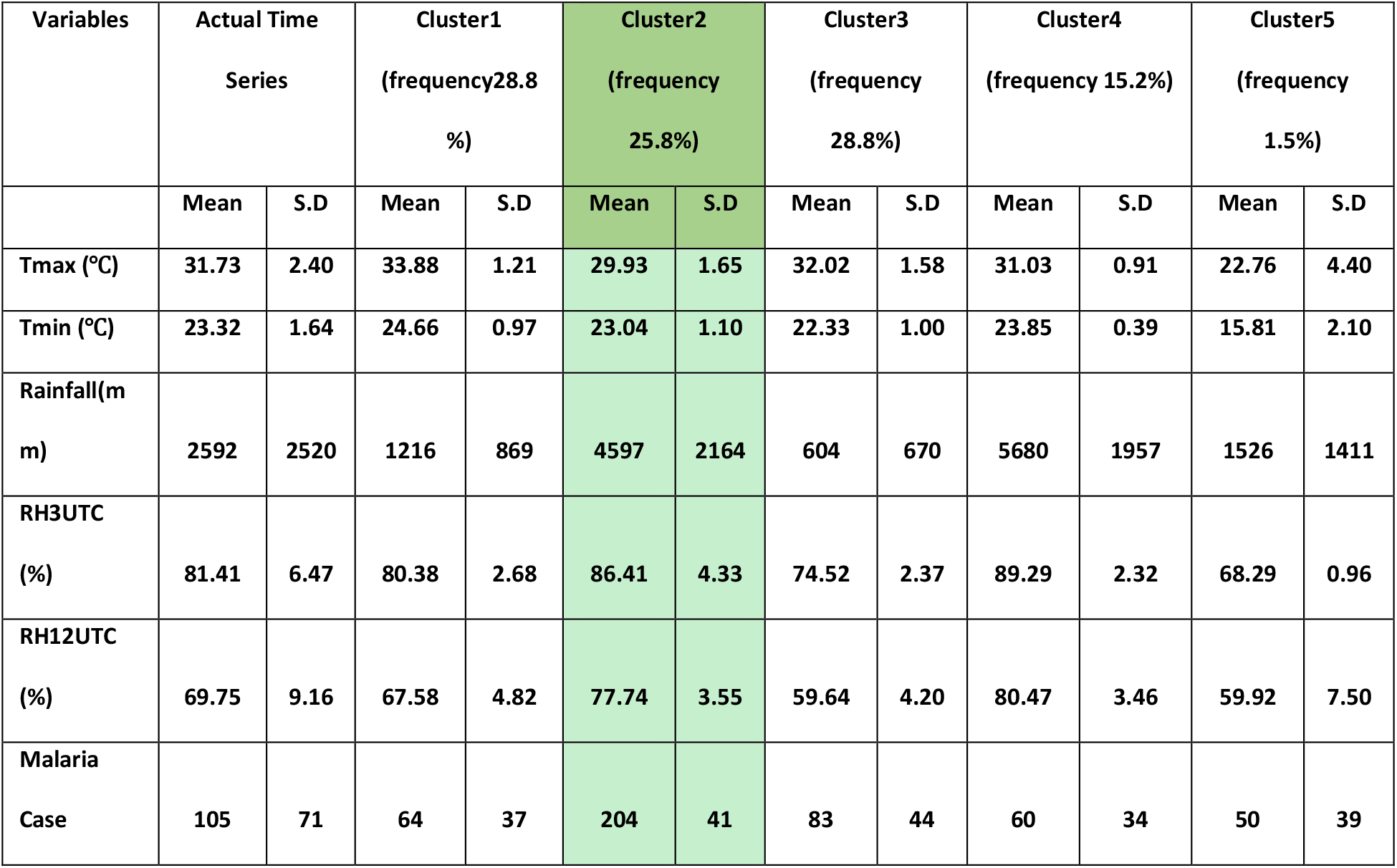
Statistics of Climate variables and Malaria cases over Kerala during 2011-2021 for actual time series and different clusters. Cluster-2 (highlighted columns) represents the malaria outbreak clusters.

**Figure 5(a-e).**
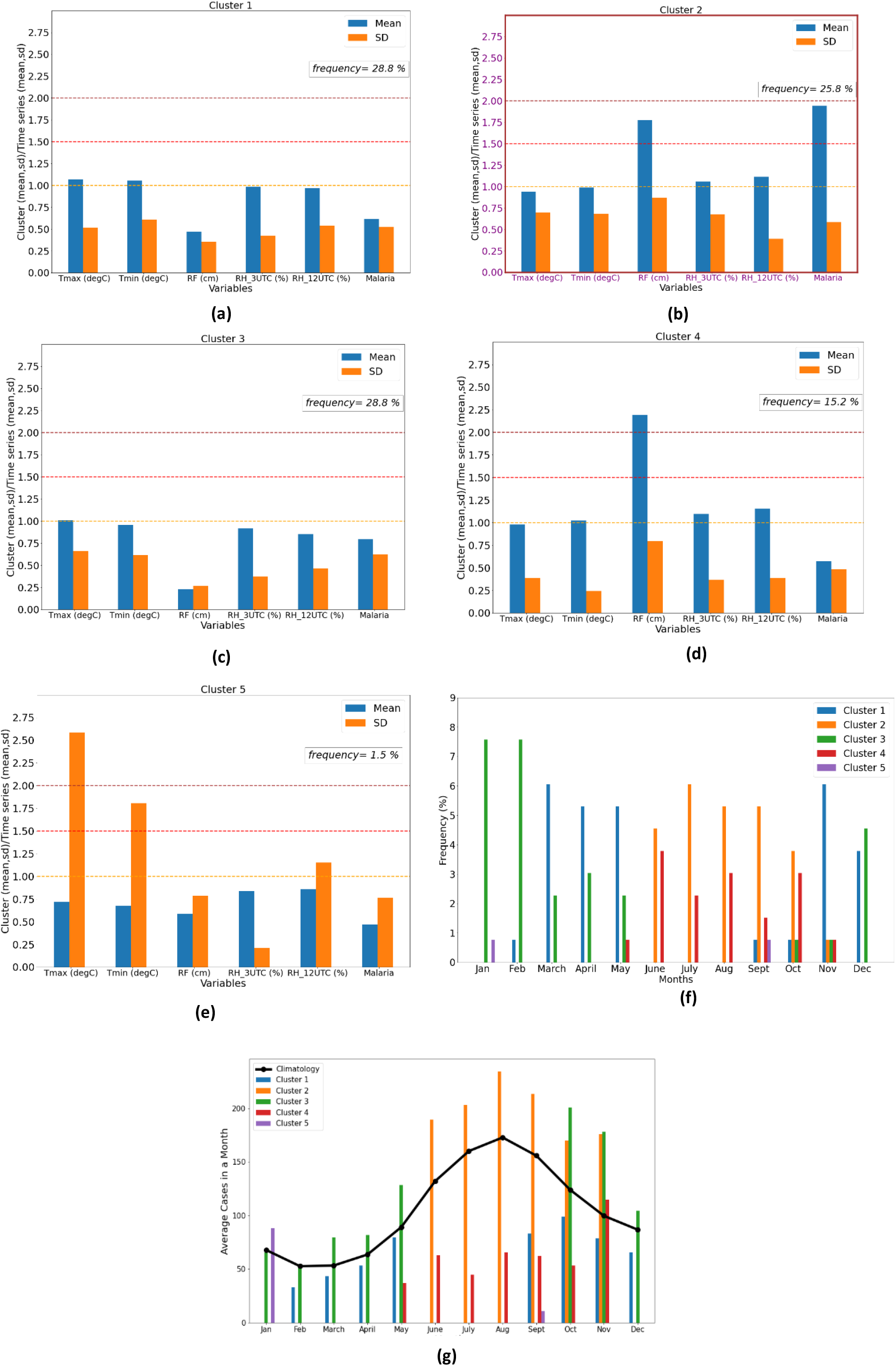
Mean and standard deviation of malaria cluster 1 to cluster 5. **(f)**. Monthly variation of malaria frequencies in cluster4. **(g)**. Monthly distribution of average malaria cases in each cluster along with malaria climatology.

**Fig.5f** shows the monthly distribution of the frequency of cases projected in each cluster. From the figure it can be seen that the cluster 2 and cluster 4 have maximum frequency projections (%) during the monsoon months (June to September). Here, there is also a seasonal splitting of cluster observed; i.e. higher values of frequencies lie in a particular season. Maximum projections of cluster 3 are seen in winter months (December, January and February) and the same for cluster 1 are in pre-monsoon (March, April and May) and post monsoon (October, November and December) months. There is a lesser chance for random projections, which means that the cluster which is projected maximum is not projected in another season.

**Fig.5g**. shows the monthly distribution of average malaria cases in each cluster along with the climatology of malaria cases. Here, it is observed that the cluster2 has maximum number of malaria cases especially in the months from June to October, which is consistent with the mean plot shown in **Fig.5b**. Similarly, in cluster4, all climate variables are near or above their time series mean, however it has reported a smaller number of Malaria cases. Here the high values of rainfall (almost twice the time series mean) and smaller standard deviation of other climate variables are not conducive for the breeding of the vectors and malaria outbreak. Earlier studies(Patz et al. 2000) ; (Kalluri et al. 2007) have shown that the increased amount of rainfall helps to washout the mosquito larva and hence reduces the chances of malaria outbreaks. So, it shows that the large number of malaria cases (204) in cluster 2 are associated with the climate conditions mentioned in the **Table 1**. Hence the fulfilment of cluster2 criteria can be considered as a threshold for malaria outbreaks.

Many times, there are some outbreaks happened in the months of October and November even though their values are less than the other months (**Fig.1d and 5g)**. So, malaria outbreaks are not completely a climate derived problem, but many times the climate factors apart from the parasite forcing can favour the disease outbreaks as the clustering analysis shows. The intensity of those outbreaks, which are directly forced by the climate factors, can be reduced by the proper climate surveillance and monitoring.

Similarly, from the cluster analysis performed for the Dengue cases (**Fig.6 (a) to (e))**, it has been observed that the cluster1 Dengue cases are approximately 1.25 times greater than the respective time series mean. The mean and standard deviation of all variables in actual time series along with that for each dengue cluster are given in **Table-2**. The specific combination of climate variables mentioned in cluster1 favour the breeding of mosquito and outbreaks of dengue cases in Kerala. It can also be noticed that the cluster 4 shows the maximum number of dengue cases (4784) however, the frequency of occurrence of cluster 4 is only 2.3% which is very much smaller than the other clusters. So, it may be considered as an outlier cluster.

**Table-2.** Statistics of Climate variables and Dengue cases over Kerala during 2011-2021 for actual time series and different clusters. Cluster-1 (highlighted columns) represents the dengue outbreak clusters.

| Variables | Actual Time Series |  | Cluster1<br>(frequency<br>27.3%) |  | Cluster2<br>(frequency<br>31.1%) |  | Cluster3<br>(frequency<br>11.4%) |  | Cluster4*<br>(frequency<br>2.3%) |  | Cluster5<br>(frequency<br>28.0%) |  |
| --- | --- | --- | --- | --- | --- | --- | --- | --- | --- | --- | --- | --- |
|  | Mean | S.D | Mean | S.D | Mean | S.D | Mean | S.D | Mean | S.D | Mean | S.D |
| Tmax (°C) | 31.73 | 2.40 | 30.79 | 0.90 | 33.62 | 1.31 | 27.39 | 2.53 | 30.68 | 0.32 | 32.40 | 1.49 |
| Tmin (°C) | 23.32 | 1.64 | 23.75 | 0.41 | 24.61 | 0.97 | 20.62 | 2.06 | 23.73 | 0.19 | 22.52 | 1.01 |
| Rainfall(m<br>m) | 2592 | 2520 | 5321 | 2301 | 1491 | 1042 | 3761 | 2108 | 4650 | 1910 | 515 | 630 |
| RH3UTC<br>(%) | 81.41 | 6.47 | 89.47 | 2.03 | 80.88 | 2.66 | 78.65 | 5.19 | 90.53 | 0.92 | 74.54 | 2.40 |
| RH12UTC<br>(%) | 69.75 | 9.16 | 80.25 | 3.15 | 69.07 | 4.84 | 70.79 | 6.35 | 80.25 | 2.93 | 59.01 | 3.78 |
| Dengue<br>Case | 485 | 782 | 668 | 487 | 343 | 428 | 289 | 193 | 4784 | 782 | 196 | 181 |

\* Cluster4 frequency is only 2.3

**Figure 6(a-e).**
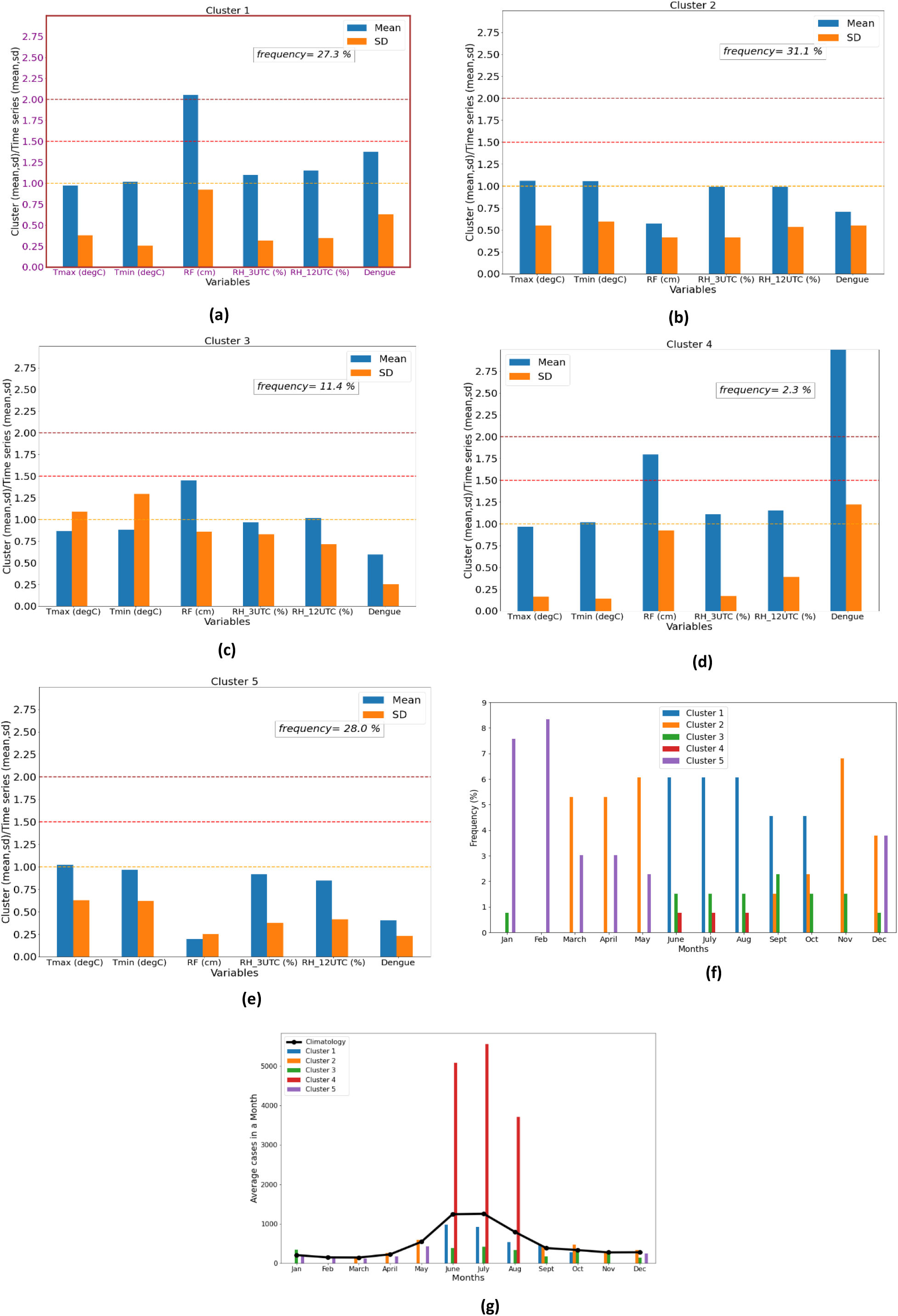
Mean and standard deviation of dengue cluster 1 to cluster 5. **(f)**. Monthly variation of dengue frequencies in cluster4. **(g)**. Monthly distribution of average dengue cases in each cluster along with dengue climatology.

The monthly frequency distribution of the projections in each cluster for dengue has shown in **Fig.6f**. It has been observed that the cluster 1 has the maximum projections during the months (June to October). Similar to Malaria clusters, here also a seasonal splitting among dengue clusters is observed. Higher values of frequencies of cluster 1 lie in the months from June to September, followed by the cluster 2 with maximum projections in the pre-monsoon (March, April and May) and post monsoon (October, November and December) months.

### (ii) Large scale climate condition results

**Fig.7(a-d)** shows the composite of climate variables (Tmax, Tmin, RH and Rainfall) over the spatial domain (10°S-50°N;40°E-120°E) in Malaria cluster 2 with a lag 0 to lag 5. In Tmax, strong positive anomalies were observed over the northern parts especially near the Tibetan plateau and negative anomalies over southern peninsular India and the northern parts of Indian Ocean. The strong negative anomalies over the southern peninsular India were observed reducing its intensity from lag5 to lag3, further becoming positive. In Tmin (**Fig.7b**), similar positive anomaly pattern was observed near the Tibetan plateau from lag 0 to lag 5. High temperature near the Tibetan anticyclone and lower temperature over the northern Indian ocean (north-south temperature gradient) are some of the large-scale climate factors which are known to impact the climate variability (Turner and Annamalai 2012),(You et al. 2020) over Indian region. Similarly, the spatial pattern of relative humidity (**Fig.7c**) and rainfall (**Fig.7d**) show high values along the west coast of India and the north-eastern region. High humidity is known to impact growth of mosquito growth (Li et al. 2013; Mohapatra et al. 2022). Hence the composite of climate factors of Malaria cluster2 are consistent with large-scale climate factors during the monsoon months.

**Figure 7:**
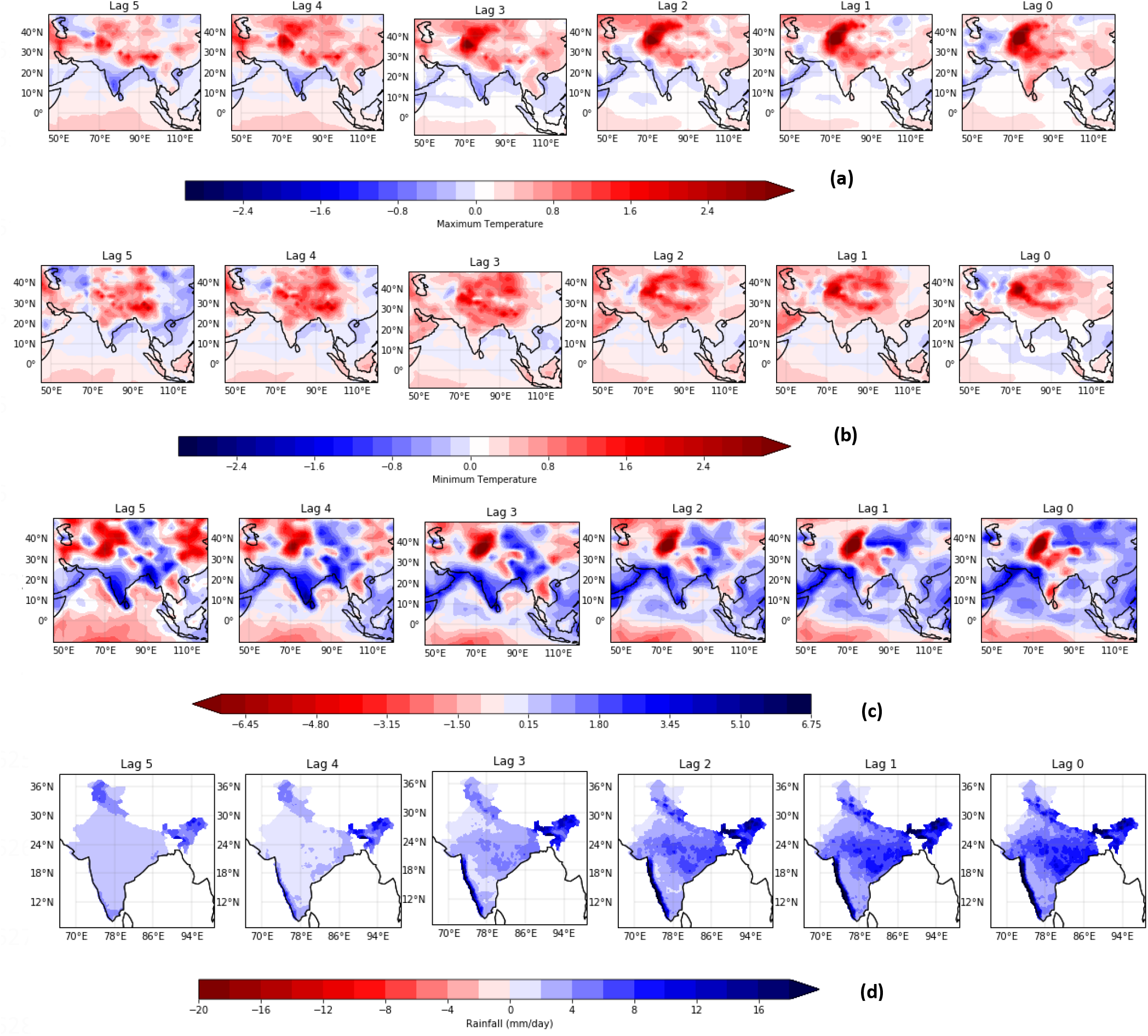
Composite of Malaria and different climate parameters for cluster2 at different lags **(a)**. Maximum Temperature **(b)**. Minimum Temperature **(c)**. Relative Humidity at 12 UTC **(d)**. Rainfall

**Fig.8(a-d)** shows the composite of climate variables (Tmax, Tmin, RH and Rainfall) in Dengue cluster 1 with a lag 0 to 5 (same spatial domain as shown for malaria cases in Fig.1). In both Tmax and Tmin, strong negative anomalies were observed over the northern parts especially near the Tibetan plateau for lag 0 to lag 5 and over peninsular India from lag 3 to lag5. Strong positive anomalies were observed for both the temperatures over the central India and the foot hills of Himalayas from lag0 to lag5. This shows the climate forcing required for the dengue outbreaks are entirely different from the forcing favouring the malaria outbreaks. The composite spatial pattern of relative humidity and rainfall of dengue cluster 1 is more or less similar to the malaria cluster2 from lag 0 to lag5. So, it shows that high amount of relative humidity and rainfall are conducive for both the disease outbreaks.

**Figure 8:**
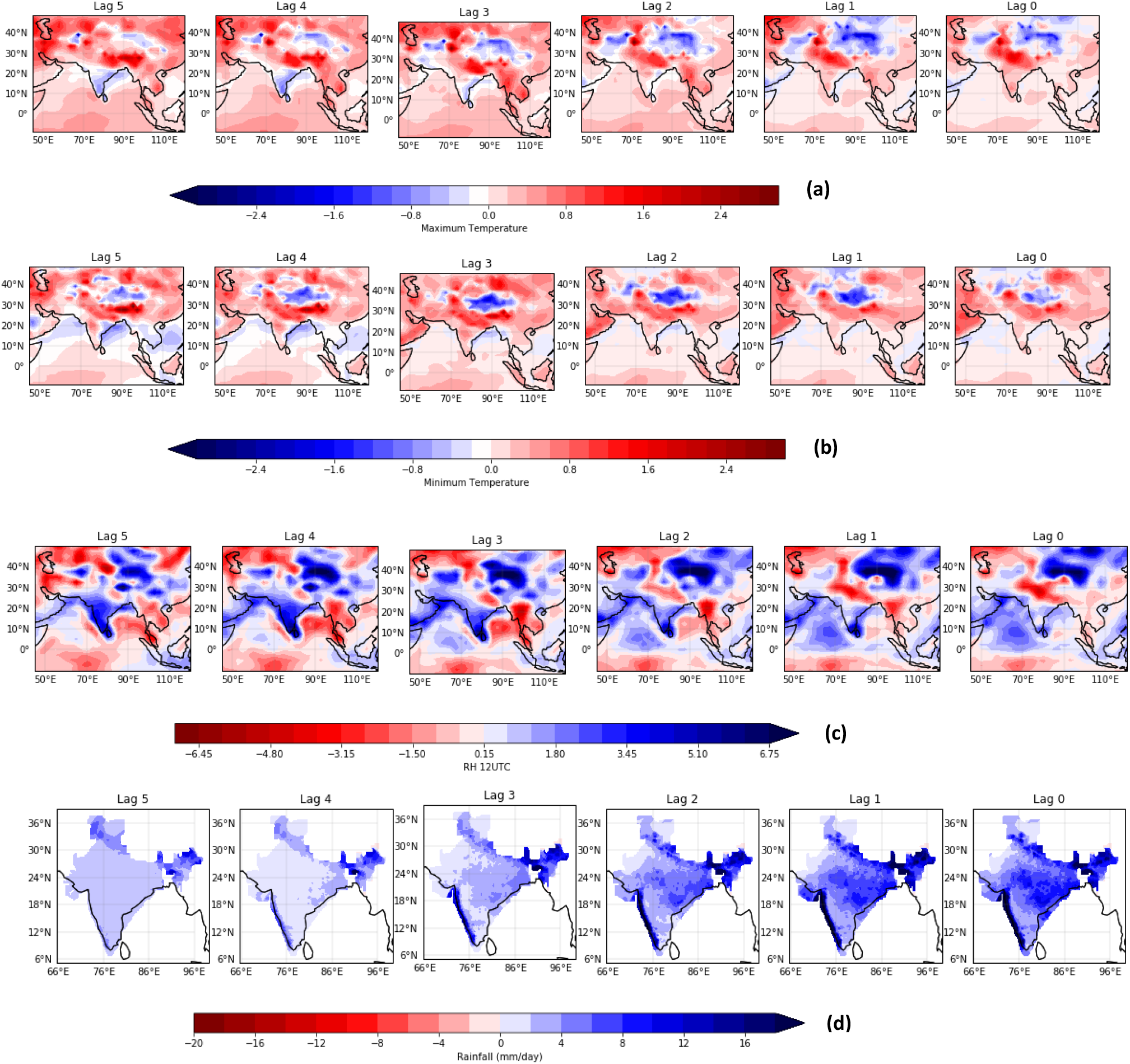
Composite of Dengue and different climate parameters for cluster2 at different lags **(a)**. Maximum Temperature **(b)**. Minimum Temperature. **(c)**. Relative Humidity at 12 UTC **(d)**. Rainfall

## Conclusion

In the current analysis we have used the malaria and dengue data over Kerala and based on a coupled system hypothesis, the outbreak of malaria and dengue are related to the multivariate clusters of climatological variables. Also, it is shown how the cyclical seasonal behaviour of the malaria and dengue cases can be explained using this model. The current analysis identified two category criteria for malaria cases (when the malaria cases are larger than the mean or when the malaria cases close to mean) which is tabulated in **Table-1**. It can be seen that high malaria cases are driven by higher than mean rainfall values and near mean values of temperature & relative humidity pattern. When the criteria is satisfied, there is a strong possibility of increase in malaria cases. Similarly, cluster analysis for dengue cases, identified one criterion (when the dengue cases are larger than the mean) which is given in **Table-2**. The dengue cases identified in this cluster are also driven by higher than mean rainfall and near mean values of temperature and relative humidity. Incidentally, an outlier cluster can be identified with large number of dengue cases but are associated with near mean values of climatic parameters (rainfall, RH etc.). However, the frequency of occurrence of this cluster is very low (only 2%).

Based on the composite analysis of climate variables, it is shown that in some cases, both malaria and dengue outbreaks over Kerala are driven by the climate forcing to some extent. However, the forcing is different for both the diseases. High temperatures over the Tibetan plateau and low temperature over the northern Indian ocean along with the high values of relative humidity and rainfall over the west coast and north-eastern parts of India are found to be allied with the malaria outbreaks over Kerala. At the same time, the out breaks of dengue are seen to be associated with the low temperatures over the Tibetan plateau and high temperatures over the central India and foot fills of Himalayas along with relative humidity and rainfall similar to that of malaria outbreaks.

The number of Malaria cases and outbreaks reported over Kerala are found to decrease in the recent years. As mentioned in the introduction, medical interventions can counteract the climate forcing and it is seen for the recent trend in malaria cases over Kerala. However, in the ongoing climate change scenario, intensity and frequency of extreme events are increasing and it may favour the suitable conditions (heavy rainfall, extreme temperatures etc.) for the outbreaks of mosquito borne diseases. In such conditions, the existing intervention methods will not be sufficient to reduce the impacts caused by the outbreaks. So, in a long-term planning perspective, along with improvements in intervention methods, implementation of the climate positive actions is also important. Coherent climate action e.g. reducing the emissions of fossil fuels and greenhouse gases, reduces the temperature extremes; Proper drainage system in a place doesn’t allow the rainwater to get stagnant over time and become conducive for the proliferation of vectors. So, these precautionary steps will not satisfy the cluster conditions which are responsible for the breeding of vector and hence reduces the frequency and intensity of MBD outbreaks. Also, the analysis suggests that, the threshold criteria has to be improved using more samples. Weekly data aggregation would improve the understanding of climate link in the control and management of MBD.

## Data Availability

All data produced in the present study are available upon reasonable request to the authors

## Acknowledgments

Authors acknowledge support from Ministry of Earth Sciences, Govt of India and the State Govt of Kerala, India for research and financial support.

## Competing Interest Statement

Authors declare that there is no competing interest

## Data Availability Statement

The disease data and the meteorological data are available in public domain. Appropriate reference is placed in the text.

## Notes

### Competing Interest Statement

The authors have declared no competing interest.

### Funding Statement

None

